# Incidence of CKD and Death among Reproductive Age Women with Dialysis Requiring Acute Kidney Injury in Ethiopia: The Role of Obstetric Risk Factors

**DOI:** 10.1101/2023.08.22.23294441

**Authors:** Ayantu Tesfaye Lemma, Tigist Workneh Leulseged, Tsion Andrias Lechebo, Sisima kornelios Osman, Delayehu Bekele Mamo

## Abstract

**Background:** Obstetric risk factors are among the leading preventable causes of Acute Kidney Injury (AKI) in hospitalized reproductive age women. Obstetric-related AKI (ORAKI) has been linked to a significant increase in the overall burden of AKI in resource-constrained settings, resulting in poor maternal and perinatal outcomes. As a result, understanding the impact of these factors on the progression of AKI is critical for a positive outcome. The study sought to determine the incidence of CKD and death, and the effect of obstetric risk factors on these outcomes among reproductive age women with dialysis requiring AKI at the national renal transplant center in Ethiopia.

**Methods:** A retrospective cohort study was conducted on 127 AKI cases (57 ORAKI and 70 None-ORAKI) who were on dialysis at the center from January 2018 to June 2020. A posthoc power analysis was calculated using G*Power 3.19.4. Data characterization and comparison was made using frequencies with percentages, median with interquartile range, chi-square test/ Fischer’s exact test and Mann-Whitney U test. A Robust Poisson regression model was used to identify factors that influence the progression of AKI to CKD and death, with Adjusted Relative Risk (ARR), 95% CIs for ARR, and P-values reported for result interpretation.

**Results:** The overall incidence rate (IR) of CKD was 5.4 per 1000 Person-days (PD) (ORAKI group=0 and None-ORAKI group= 9.7 per 1000 PD) and the overall incidence rate of death was 7.8 per 1000 PD (ORAKI group=5.5 per 1000 PD and None-ORAKI group= 9.7 per 1000 PD). According to the multivariable regression analysis, participants with ORAKI had a 22% lower risk of progression to CKD or death than those with None-ORAKI (ARR=0.78, 95%CI=0.67-0.90, p=0.001).

**Conclusions:** Although having obstetric related risk factors has been linked to an increased risk of developing AKI, once it occurs, those with ORAKI have a significantly better prognosis than those with None-ORAKI. Continued efforts to prevent AKI in pregnant women and to slow its progression once it has developed are critical for a better maternal and fetal outcome.

## INTRODUCTION

Acute kidney injury (AKI) refers to an abrupt (within hours) decrease in kidney function, resulting in the retention of urea and other nitrogenous waste products that, unless treated early, can result in a number of systematic complications, end stage renal failure and death (1,2). AKI is caused by a variety of factors, including high-risk patient characteristics and other external insults that can damage the kidney via various mechanisms (3-19). The outcome can range from full renal recovery to progression to CKD and death, depending on the severity of the underlying condition and the timing of treatment initiation (19-26).

Obstetric risk factors are among the leading preventable causes of Acute Kidney Injury (AKI) in hospitalized reproductive age women. Obstetric related AKI (ORAKI) has been reported to contribute significantly to the overall burden of AKI in resource-limited settings, resulting in poor maternal and fetal outcomes (21,25,27). Given that the majority of obstetric-related maternal deaths occur in developing countries, particularly Sub-Saharan Africa, understanding the major causes, such as AKI, is critical because these are conditions that can be avoided and treated with a good prognosis for both the mother and fetus if detected early (27).

Pre-eclampsia and eclampsia were found to be the most common causes of ORAKI in any setting, accounting for up to three-quarters of cases. In developing countries, this is followed by puerperal sepsis and shock, both of which account for a sizable proportion (28-32). According to reports, the prognosis for ORAKI is good, with a large proportion of these cases, 50 to 80 percent, recovering from AKI. In the remaining one-fifth to one-quarter, they either progressed to end-stage renal disease or died (28,29,33).

So far, studies on ORAKI have focused on characterizing patients and determining its effect on the development of AKI, demonstrating that it is a significant factor and that the risk is further determined by other personal and medical conditions (32,34). However, there has been little research done to assess the effect of obstetric risk factors on the progression of AKI, once it occurred, and its outcome, particularly in developing countries. Hence, the objective of this study is to determine the incidence of CKD and death, and the effect of obstetric risk factors on these outcomes among reproductive age women with dialysis requiring AKI at the national renal transplant center in Ethiopia from January 2018 to June 2020.

## METHODOLOGY

### Study Design, Population and Sample Size

An institution based retrospective cohort study was conducted from October to November, 2020 at St Paul’s Hospital Millennium Medical College (SPHMMC), a tertiary teaching hospital under the Federal Ministry of Health in Addis Ababa, Ethiopia. SPHMMC is the country’s second largest governmental hospital, as well as the first governmental hospital to provide both acute and chronic hemodialysis services in collaboration with the Egyptian government, and thus serves as the primary government-owned referral center for AKI patients. The hospital currently has around forty hemodialysis machines for end-stage renal disease and six hemodialysis machines for acute kidney injury.

The exposure groups are as follows:

- Women of reproductive age (15-49 years) with AKI 2° to obstetric risk factors/ ORAKI (**Exposed group**): These are cases who were admitted to the center for dialysis between January 2018 to June 2020 for AKI following obstetric related risk factors.
- Women of reproductive age (15-49 years) with AKI 2° to none obstetric risk factors/None-ORAKI (**Non-exposed group**): These are cases who were admitted to the center for dialysis between January 2018 to June 2020 for AKI due to causes other than obstetric related risk factors.

During the observation period, a total of 143 women were managed at the center. Of which, 127 were in the reproductive age group and were included in the study. Among the 127 cases, 57 were women with AKI 2° to obstetric risk factors and 70 were women with AKI 2° to none obstetric risk factors. **(Figure 1)**

**Figure 1:**
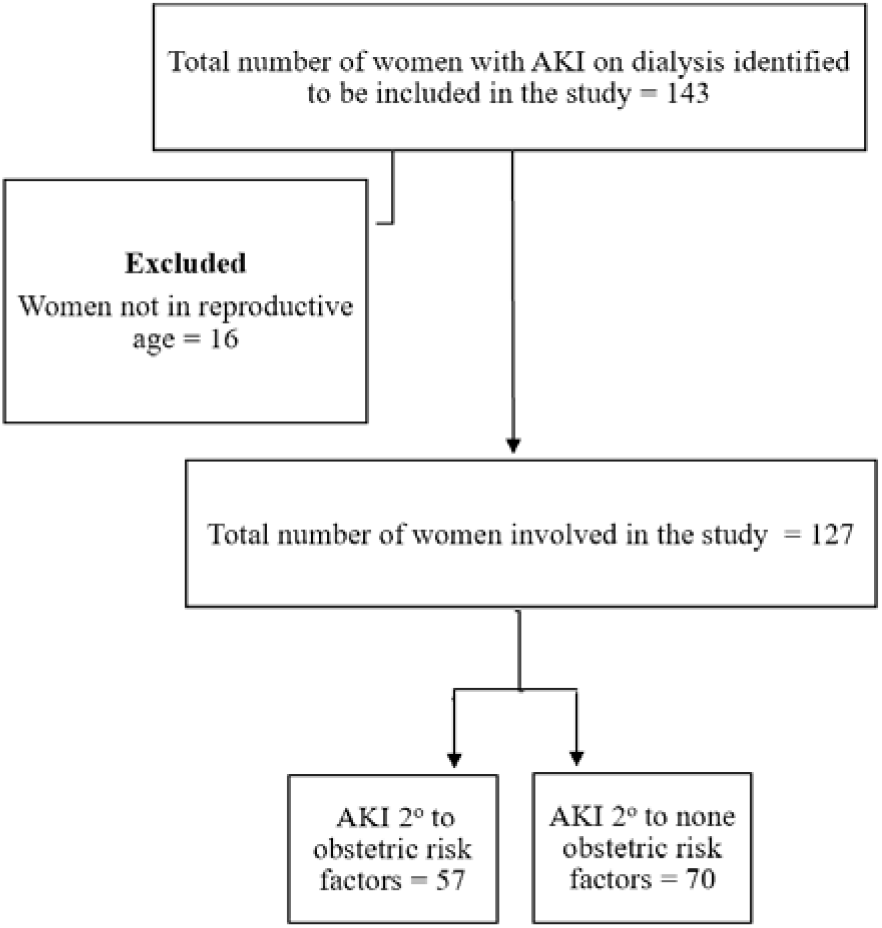
Flow chart showing the disposition of study participants in the final analysis

A post-hoc power analysis was calculated using G*Power 3.19.4 to check the power of the study using a two-tailed z-test for difference between two independent proportions with the following statistical parameters; 5% level of significance, prevalence and sample size in the None-ORAKI group of 41.3% and 63, respectively; and prevalence and sample size in the ORAKI group of 12.0% and 50, respectively. Finally, the power of the study was found to be 94.5%.

### Operational Definition

**Progression to chronic kidney disease**: It is diagnosed when a patient who is initially admitted with an assessment of AKI and kept on hemodialysis failed to show clinical or biochemical improvement and later diagnosed to have CKD.

### Data collection and Quality assurance

A pretested data abstraction tool was used to extract data from patients’ charts. The tool included questions on socio-demographics, medical illness history, obstetric and surgical history, exposure to nephrotoxic drugs, pre-renal causes, and AKI outcome. Two trained General Practitioners extracted the data. To ensure the data’s quality, it was cleaned by checking for inconsistencies, numerical errors, and missing parameters, and appropriate measures were taken. In the event of a discrepancy in patient information, data was double-checked by referring to the main record and cross-referencing with another database whenever possible. SPSS version 25.0 software was used for all data management and analysis.

### Statistical analysis

To characterize the study population, descriptive analysis using frequencies with percentages and median with interquartile range (after testing the assumption of normality) was run. To measure the outcome, incidence density with 95% CI was run.

A chi-square test/ Fischer’s exact test and Mann-Whitney U test were used to identify the presence of a statistically significant difference between the ORAKI and None-ORAKI groups in terms of their underlying characteristics. A statistically significant difference was detected for variables with a p-value of ≤ 0.05.

To identify the effect of obstetric related risk factors on outcome of AKI, Robust Poisson regression model was used. Univariate analysis at 25% level of significance was run to identify exposures to be controlled for in the final analysis. On the final multivariable analysis, at 5% level of significance, adjusted RR, P-value and 95% CI for RR were used to test significance and interpretation of results. Variables with p-value ≤ 0.05 were considered as significant predictors of AKI outcome.

## RESULTS

### Baseline characteristics of participants

From the 127 participants, majority reside in two regions; 52 (40.9%) in Oromia and 24 (18.9%) in Addis Ababa, and were between the ages of 25 and 34 (38.6%). One or more underlying acute and/or chronic medical illnesses were present in 96 (75.6%) participants. Hypertension and cardiovascular disease were the most common chronic illnesses, accounting for 33 (26.0%) and 15 (11.8%) of all cases, respectively. The most common acute conditions were sepsis in 52 (40.9%) and shock in 36 (28.3%). AKI caused by pregnancy was diagnosed in 57 (44.9%). From these 57 patients, 50 had preeclampsia/HELLP syndrome, 14 had peripartum hemorrhage, and 8 had puerperal sepsis, with the majority having more than one complication, particularly preeclampsia/HELLP syndrome with the other two. The vast majority (94.5%) had a history of exposure to at least one nephrotoxic drug. PPI (110/120), vancomycin (67/120), and ceftriaxone (44/120) were the most commonly used medications. Glomerular disease was found in 91 (71.7%) of the participants. ATN was the most common type (56/91), with 26/56 being ischemic ATN and the remaining 30/56 being septic ATN. AGN (18/91), RPGN (18/91), and PIGN (11/91) were also diagnosed in the majority of cases. Only two (1.8%) cases had a post-renal cause, both of which were due to ureteric stone. Major surgery was performed on 23 (18.1%) of the participants for obstetric or other reasons. Twenty-three (18.1%) of the participants required ICU admission, with 14 (24.6%) having a pregnancy-related risk factor and 9 (2.9%) having no such risk.

A statistically significant difference in age, exposure to vancomycin, ACEIs/ARBs, and PPI, diagnosis of AGN and RPGN, and history of major surgery was found when the underlying characteristics of participants were compared between groups with ORAKI and None-ORAKI. Accordingly, a significantly higher proportion of participants with ORAKI were younger, mainly 25-34 years (50.9%), than those with None-ORAKI who were mostly 35-49 years of age (41.4%) (p=0.007). In terms of drug exposure, vancomycin was taken by a greater proportion of participants with ORAKI (66.7% vs 41.4%, p=0.005). On the other hand, a higher proportion of participants with None-ORAKI were exposed to PPI (77.2% vs 94.3%). Furthermore, 7 (10%) of participants with None-ORAKI took ACEIs/ARBs, with a p-value of 0.016 indicating a significant difference between the two groups. AGN (5.3% vs 21.4%, p=0.009) and RPGN (5.3% vs 21.4%, p=0.009) were diagnosed at a significantly lower proportion in the group with ORAKI. Finally, a significantly higher proportion of participants with ORAKI (28.1% vs 10.0%, p=0.009) underwent major surgery. **(Table 1)**

**Table 1:** Baseline characteristics of participants and comparison based on exposure status (n=127)

| Variable |  | ORAKI |  | Total (%) | p-value |
| --- | --- | --- | --- | --- | --- |
|  |  | Yes (n=57) | No (n=70) |  |  |
| *Age category (in years)* | 15-24 | 18 (31.6) | 21 (30.0) | 39 (30.7) | **0.007\*** |
|  | 25-34 | 29 (50.9) | 20 (28.6) | 49 (38.6) |  |
|  | 35-49 | 10 (17.5) | 29 (41.4) | 39 (30.7) |  |
| *Hypertension* | Yes | 14 (24.6) | 19 (27.1) | 33 (26.0) | 0.741 |
|  | No | 43 (75.4) | 51 (72.9) | 94 (74.0) |  |
| *Cardiovascular illness* | Yes | 4 (7.0) | 11 (15.7) | 15 (11.8) | 0.131 |
|  | No | 53 (93.0) | 59 (84.3) | 112 (88.2) |  |
| *Diabetes mellitus* | Yes | 0 | 4 (5.7) | 4 (3.1) | 0.127 |
|  | No | 57 (100.0) | 66 (94.3) | 123 (96.9) |  |
| *HIV* | Yes | 2 (3.5) | 5 (7.1) | 7 (5.5) | 0.458 |
|  | No | 55 (96.5) | 65 (92.9) | 120 (94.5) |  |
| *Sepsis* | Yes | 24 (42.1) | 28 (40.0) | 52 (40.9) | 0.810 |
|  | No | 33 (57.9) | 42 (60.0) | 75 (59.1) |  |
| *Shock* | Yes | 16 (28.1) | 20 (28.6) | 36 (28.6) | 0.950 |
|  | No | 41 (71.9) | 50 (71.4) | 91 (71.7) |  |
| *Vancomycin* | Yes | 38 (66.7) | 29 (41.4) | 67 (52.8) | **0.005\*** |
|  | No | 19 (33.3) | 41 (58.6) | 60 (47.2) |  |
| *Ceftriaxone* | Yes | 19 (33.3) | 25 (35.7) | 44 (34.6) | 0.779 |
|  | No | 38 (66.7) | 45 (64.3) | 83 (65.4) |  |
| *ACEIs/ARBs* | Yes | 0 | 7 (10.0) | 7 (5.5) | **0.016\*** |
|  | No | 57 (100.0) | 63 (90.0) | 120 (94.5) |  |
| *NSAIDs* | Yes | 0 | 3 (4.3) | 3 (2.4) | 0.252 |
|  | No | 57 (100.0) | 67 (95.7) | 123 (97.6) |  |
| *PPI* | Yes | 44 (77.2) | 66 (94.3) | 110 (86.6) | **0.005\*** |
|  | No | 13 (22.8) | 4 (5.7) | 17 (13.4) |  |
| *AGN* | Yes | 3 (5.3) | 15 (21.4) | 18 (14.2) | **0.009\*** |
|  | No | 54 (94.7) | 55 (78.6) | 109 (85.8) |  |
| *RPGN* | Yes | 3 (5.3) | 15 (21.4) | 18 (14.2) | **0.009\*** |
|  | No | 54 (94.7) | 55 (78.6) | 109 (85.8) |  |
| *PIGN* | Yes | 6 (16.2) | 5 (7.1) | 11 (10.3) | 0.184 |
|  | No | 31 (83.8) | 65 (92.9) | 96 (89.7) |  |
| *ATN* | Yes | 27 (47.4) | 29 (41.4) | 56 (44.1) | 0.502 |
|  | No | 30 (52.6) | 41 (58.6) | 71 (55.0) |  |
| *Ureteric stone* | Yes | 1 (1.8) | 1 (1.4) | 2 (1.6) | 1.000 |
|  | No | 56 (98.2) | 69 (98.6) | 125 (98.4) |  |
| *Major surgery* | Yes | 16 (28.1) | 7 (10.0) | 23 (18.1) | **0.009\*** |
|  | No | 41 (71.9) | 63 (90.0) | 104 (81.9) |  |
| *ICU admission* | Yes | 14 (24.6) | 9 (12.9) | 23 (18.1) | 0.088 |
|  | No | 43 (75.4) | 61 (87.1) | 104 (81.9) |  |

### Total and Sub-group Incidence Density of CKD and Death

Among the 127 patients, outcome data was not recorded for 14 participants who were transferred to another facility for reasons other than medical indications. A statistical comparison of the underlying characteristics of the 14 transferred patients and the remaining 113 patients was performed to determine the presence of significant differences in their exposure to important factors that could have made them more inclined to be at high risk of developing one group of outcomes, potentially biassing the overall result of the study findings. As a result, no significant difference was found in any of the comparisons (p-values for chi-square/Fischer’s exact tests and Mann-Whitney U-tests were greater than 0.05).

The remaining 113 participants were followed for a median of 21.0 days (IQR, 13.0-25.0), and there was no significant difference in follow-up duration between the two groups (p=0.938). The overall incidence rate (IR) of CKD was 5.4 per 1000 person-days (PD) of observation (95% CI=3.1 - 9.3). The overall death rate was 7.8 per 1000 PD (95% CI= 5.0 - 12.3). According to the sub-group analysis, none of the cases with ORAKI progressed to CKD, and six died, resulting in a death rate of 5.5 per 1000 PD (95% CI= 2.5-12.3). Among those with None-ORAKI, 13 developed CKD (IR=9.7 per 1000 PD observation, 95% CI=5.7-16.8) and 13 died (IR=9.7 per 1000 PD observation, 95% CI=5.7-16.8). **(Table 2)**

**Table 2:** Comparison of AKI outcomes and length of stay based on exposure status (n=113)

| Variable | ORAKI |  | Total (%) |
| --- | --- | --- | --- |
|  | Yes (n=50) | No (n=63) |  |
| **Disease outcome (n, %)** |  |  |  |
| Progression to CKD | 0 | 9.7 per 1000 PD | 5.4 per 1000 PD |
| Death | 5.5 per 1000 PD | 9.7 per 1000 PD | 7.8 per 1000 PD |
| **Length of stay in days (Median, IQR)** | 21.0 (14.8-21.5) | 20.0 (13.0-26.0) | 21.0 (13.0-25.5) |

### Effect of obstetric related risk factor on AKI outcome

To assess the effect of obstetric related risk factor on AKI outcome, a multivariable Robust Poisson Regression model was run after adjusting for age category, pregnancy related risk factor, cardiovascular disease, vancomycin, ceftriaxone, PPI, AGN, PIGN, RPGN, ATN and surgery which were found to be significantly associated with AKI outcomes on the univariate analysis.

Accordingly, the risk of progression to CKD or death among participants with ORAKI was 22% lower than those with None-ORAKI (ARR=0.78, 95%CI=0.67-0.90, p=0.001).

In addition, cardiovascular disease and taking vancomycin were found to be significantly associated with AKI outcome. Having cardiovascular disease and taking vancomycin were associated with an increased risk of progression to CKD and death by 18% (ARR=1.18, 95%CI=1.01,1.39, p=0.044) and 19% (ARR=1.19, 95%CI=1.05,1.34, p=0.006) as compared to those with no cardiovascular disease and didn’t take vancomycin, respectively. (**Table 3**)

**Table 3:**
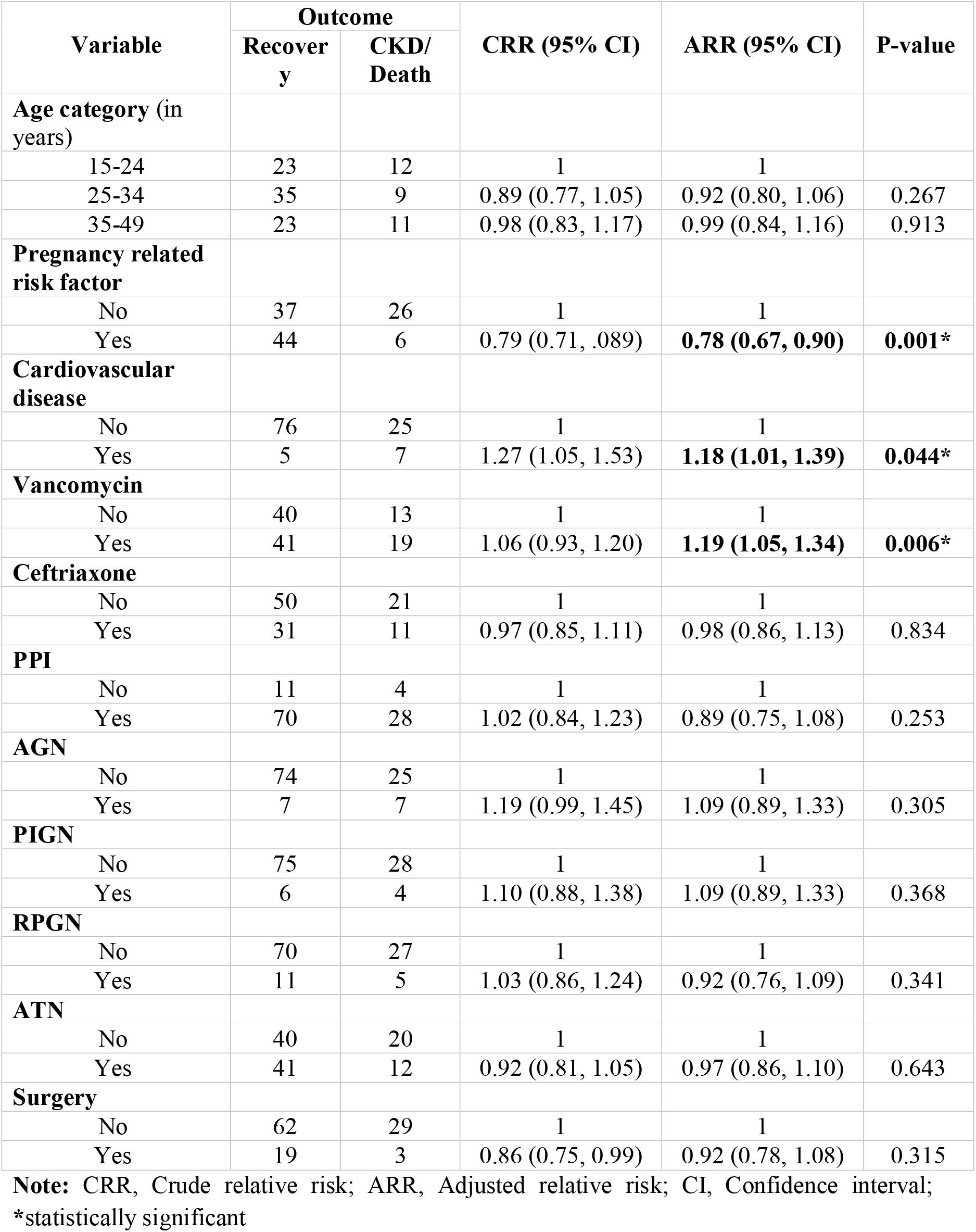
Predictors of AKI outcome among patients on dialysis (n=113)

## DISCUSSION

Among the 127 participants, ORAKI was diagnosed in 57 (44.9%). From which, 50/57 had preeclampsia/HELLP syndrome, 14/57 had peripartum hemorrhage and 8/57 had puerperal sepsis. This pattern corresponds to the most common cause of obstetric-related AKI in both developed and developing countries (28-32).

The overall incidence rate (IR) of CKD was 5.4 per 1000 PD (ORAKI = 0 and None-ORAKI= 9.7 per 1000 PD) and the overall incidence rate of death was 7.8 per 1000 PD (ORAKI=5.5 per 1000 PD and None-ORAKI=9.7 per 1000 PD). This demonstrates that the majority of ORAKI cases recovered when compared to those with None-ORAKI. Such favorable outcome is also reported in other studies conducted in Ethiopia, Tanzania and Iran, where a higher proportion of cases with obstetric related causes recovered and a lower proportion progressed to CKD and died (28,29,30). Further regression analysis to determine the effect of obstetric factors on AKI outcome revealed that cases with ORAKI had a 22% lower risk of progressing to CKD or dying, indicating that cases with ORAKI have a much better prognosis than cases with Non-ORAKI. This could be because most ORAKI cases are otherwise healthy, with few underlying medical conditions that could lead to complication, as in our study, where a significantly higher proportion had clinically favorable factors such as younger age, less frequent underlying glomerular disease, particularly AGN and RPGN, and lack of exposure to specific nephrotoxic drugs such as ACEIs/ARBs. Moreover, because ORAKI occurs following pregnancy, labor and delivery while the woman is under strict medical care, timely diagnosis of AKI and initiation of management is highly likely, contributing to the favorable outcome.

Furthermore, cardiovascular disease and vancomycin exposure were linked to an 18% and 19% increased risk of CKD progression and death, respectively. These factors have also been shown to be significant predictors in a number of other studies, owing to their underlying pathophysiologic mechanisms that damage the kidney and their prevalence/exposure in the majority of cases (16,24,25).

The study’s finding is a good addition to the existing literature because it addressed a less studied research question, was conducted in the largest national referral dialysis Centre, and had an adequate sample size with a post-hoc achieved power of 94.5%. As a result, the obtained result is sufficiently powered. However, because the study was conducted retrospectively and there was insufficient chart recording with limited information on additional exposures, further relevant confounders were not controlled for.

## CONCLUSIONS

Although having obstetric related risk factors has been reported to be associated with an increased risk of developing AKI, once it occurs, the prognosis for those with ORAKI with standard management is significantly better than those with None-ORAKI. Continued efforts to prevent the development of AKI in pregnant women and minimize its progression once it has occurred are essential for a better maternal and fetal outcome. In addition, stringent monitoring of those with cardiovascular disease and those taking vancomycin is crucial to mitigate risk of progression. To generate additional evidence, a large-scale cohort study with a thorough assessment of all exposures is required.

## Data Availability

All data produced in the present study are available upon reasonable request to the authors

## List of Abbreviations

ACEIs/ARBs: Angiotensin-Converting Enzyme Inhibitors/ Angiotensin II Receptor Antagonists
AKI: Acute kidney injury
ATN: Acute Tubular Necrosis
AGN: Acute Glomerulonephritis
CI: Confidence interval
Cr: Creatinine
HELLP: hemolysis, elevated liver enzymes, and low platelets
HIV: Human immunodeficiency virus
IRB: Institutional review board
ICU: Intensive Care Unit
NSAIDs: Non-steroidal anti-inflammatory drugs
ORAKI: Obstetric Related Acute Kidney Injury
RR: Relative Risk
PIGN: Post-infectious Glomerulonephritis
PPI: Proton Pump Inhibitors
RPGN: Rapidly Progressive Glomerulonephritis

## Declaration

### Ethics approval

The study was conducted after securing ethical clearance from St. Paul’s Hospital Millennium Medical College institutional review board (SPHMMC-IRB). The St. Paul’s Hospital Millennium Medical College institutional review board (SPHMMC-IRB) also waived the need for informed consent since the study used secondary data (IRB letter no: PM.23/724). The study was carried out in accordance with relevant guidelines and regulations. Medical record number was used for the data collection and personal identifiers of the patient were not used in the research report. Access to the collected information was limited to the research team and confidentiality was maintained throughout the project.

### Consent to participate

Not applicable

### Availability of data and materials

All relevant data are available upon reasonable request from Tigist Workneh Leulseged at.

### Author’s Contribution

ATL and TWL conceived and designed the study. DBM, TAL, and SKO contributed to the conception and design of the study. ATL supervised the data collection. TWL performed statistical analysis, and drafted the initial manuscript. TAL and SKO contributed to statistical analysis. DBM revised the draft manuscript. All authors approved the final version of the manuscript.

## Acknowledgement

The authors would like to thank St. Paul’s Hospital Millennium Medical College for sponsoring and facilitating the research work.

